# Efficacy of bladder training in adults with overactive bladder: A systematic review protocol

**DOI:** 10.1101/2022.03.01.22271687

**Authors:** Anna Karoline Lopes Rocha, Silvia Elizate Monteiro, Maria P. Volpato, Dinah Verleun de Paula e Silva, Lilian Valim, Cássio Riccetto, Simone Botelho

## Abstract

**Introduction:** The aim of this systematic review will be to investigate and update whether bladder training can promote improvement of symptoms of overactive bladder syndrome with or without urgency urinary incontinence in adults.

**Methods:** We will perform a systematic review according to the Cochrane methodology of randomized controlled trials. An overall search strategy will be developed and adapted for each database. A bibliographic search will be conducted in eight databases - PubMed, PEDro, SciELO, LILACS, Cochrane Library, Web of Science, EMBASE, CINAHL, by manual searching. The MeSH terms will be “Bladder Training”, “Bladder Drill”, “Bladder Re-education”, “Bladder Retraining”, “Bladder Discipline”, “Overactive Bladder”, “Bladder, Overactive”, “Overactive Urinary Bladder”, “Urinary Bladder”, “Overactive, Urinary Bladder”, “Bladder, Urinary”, “Urinary Bladder Disease”, “Bladder Disease”, “Bladder Detrusor Muscle” and “Detrusor Muscle, Bladder”. Meta-analysis, if plausible, will be performed by the software Review Manager 5.4. *Primary outcomes*: symptoms of overactive bladder syndrome (urinary urgency with daytime voiding frequency, nocturia with or without urgency urinary incontinence), and cure/improvement assessed by instruments. *Secondary outcomes*: quality of life, functional assessment, validated scale/questionnaire and adverse events. Quality assessment will be performed by Cochrane instrument and quality of evidence will be assessed by GRADE.

**Discussion:** This study is a review of randomized controlled studies to analyze the efficacy of bladder training in improving the symptoms of adults with overactive bladder syndrome. The study design of randomized controlled trials for a higher level of scientific evidence was chosen. The aim is to obtain results that allow further studies and evidence that this intervention generates beneficial effects in the sample studied.

**Trial registration:** 

**Systematic review registration:** PROSPERO CRD42022301522.

## Introduction

According to the International Continence Society (ICS), the overactive bladder (OAB) is a multifactorial clinical syndrome associated with intrinsic and extrinsic factors, and is defined as the presence of symptoms of urinary urgency, daytime voiding frequency and/or nocturia, with urinary incontinence (OAB-wet) or without (OAB-dry), in the absence of urinary tract infection or other detectable disease [1-6]. The OAB tends to directly impact the quality of life of individuals, affecting self-esteem and interpersonal relationships, and contributing to social isolation [7,8]. However, the search for treatment is usually delayed due to embarrassment and lack of knowledge [9].

Its prevalence is quite variable (2 to 53%), ranging from 2 to 35% among men and 3 to 41% among women, tending to increase with age in both genders [8,10,11,12]. The higher prevalence of the female population may be related to different factors, which was suggested by Peyronnet *et al*. (2019), which highlights the relevance of phenotypes for a better understanding of the trigger mechanisms and the importance of the individualized approach [13].

On the other hand, OAB is a dynamic condition, with a remission rate of 49% in OAB-dry and 26% in OAB-wet women, which tends to be costly for public health.

According to international guidelines, low-cost and low-complexity therapeutic strategies are recommended as first-line treatment, aiming to promote a good cost-benefit relationship as well as quality of life in individuals with OAB [3,5,7,8,14-18].

The first-line conservative approach to the symptoms of OAB-wet or OAB-dry includes behavioral interventions, which consists of strategies that modify lifestyle, life habits, and patient environment, with scheduled voiding regimens, including bladder training (BT) and pelvic floor muscle training (PFMT) [16-19]. BT has been shown to be important, not only for the results presented, but because it has low cost, low complexity and reduced side effects [20].

BT consists of a set of techniques that help individuals to delay voiding through activities that require mental concentration, such as relaxation or distraction techniques, often associated with repeated contractions of pelvic floor muscles (PFM), which provide an inhibitory reflex of the detrusor (Urethrosphincteric guarding reflex) [18, 21, 22]. Therefore, while the individual to delay voiding, the voluntary contractions of PFM activate afferent stimulus, via the pudendal nerve to the sacral center of the urination with inhibitory responses to the detrusor through the pelvic nerve (Perineodetrusor inhibitory reflex), resulting in increased intervals between the voidings [22].

The review published was study observed that are limited evidence available, what suggests that BT may be helpful for the treatment of individuals with symptoms of OAB, but there was also not enough evidence to determine whether BT was useful as a supplement to another therapy, either by the quality of evidence of the studies or by the sample size, confidence interval or estimates of effect [23].

Considering this context, the aim of this study is to investigate the effect of BT on the symptoms of OAB-wet or OAB-dry in adults, through a systematic review of available randomized clinical trials (RCTs).

## Methods

### Systematic review

#### Study inclusion and exclusion criteria

##### Inclusion Criteria

RCTs of adults over 18 years old with OAB, with or without urgency urinary incontinence, receiving any kind of BT intervention with or without supervision, to prevent and/or reduce the symptoms of OAB.

##### Exclusion criteria

Case studies, cross-over studies, gray literature studies, or those that do not have the separation of the groups nor details of the adequate protocol.

### Outcomes

#### Primary outcomes

*C*ure/improvement of the OAB symptoms (urinary urgency with daytime voiding frequency, nocturia with or without urgency urinary incontinence) assessed by quantity instruments.

#### Secondary outcomes

Quality of life, functional assessment and adverse events.

### Literature search strategies

A way strategy will be created and adapted for searching the databases PubMed, Physiotherapy Evidence Database (PEDro), Scientific Electronic Library Online (SciELO), Latin American and Caribbean Literature in Health Sciences (LILACS), Central Cochrane Library, Web of Science, EMBASE, CINAHL (Cummulative Index to Nursing and Allied Health Literature), and by manual searching to identify studies involving the above-mentioned interventions. Last date of search will until December 2022. There will be no language restrictions. MeSH terms that will be used for our searches are “Bladder Training”, “Bladder Drill”, “Bladder Re-education”, “Bladder Retraining”, “Bladder Discipline”, “Overactive Bladder”, “Bladder, Overactive”, “Overactive Urinary Bladder”, “Urinary Bladder”, “Overactive, Urinary Bladder”, “Bladder, Urinary”, “Urinary Bladder Disease”, “Bladder Disease”, “Bladder Detrusor Muscle” and “Detrusor Muscle, Bladder”.

### Data collection and analysis

#### Selection of studies

At this study, for each search strategy, two reviewers will independently evaluate the studies gathered from the databases in the order: title, abstract and full reading. All studies potentially eligible for inclusion in the review will be selected for full reading. In case of disagreement, a third reviewer will be consulted.

### Data extraction and management

Data extraction for eligible studies will be performed by two reviewers (AKLR e DPVS) who will independently extract data from articles that meet the inclusion criteria. A standardized form will be used to extract the following information: study characteristics (design, randomization method, blinding, allocation generation and concealment, statistics); participants; interventions; clinical outcomes (types of outcomes measured: dichotomous or continuous as shown in Table 1.

**Table 1.**
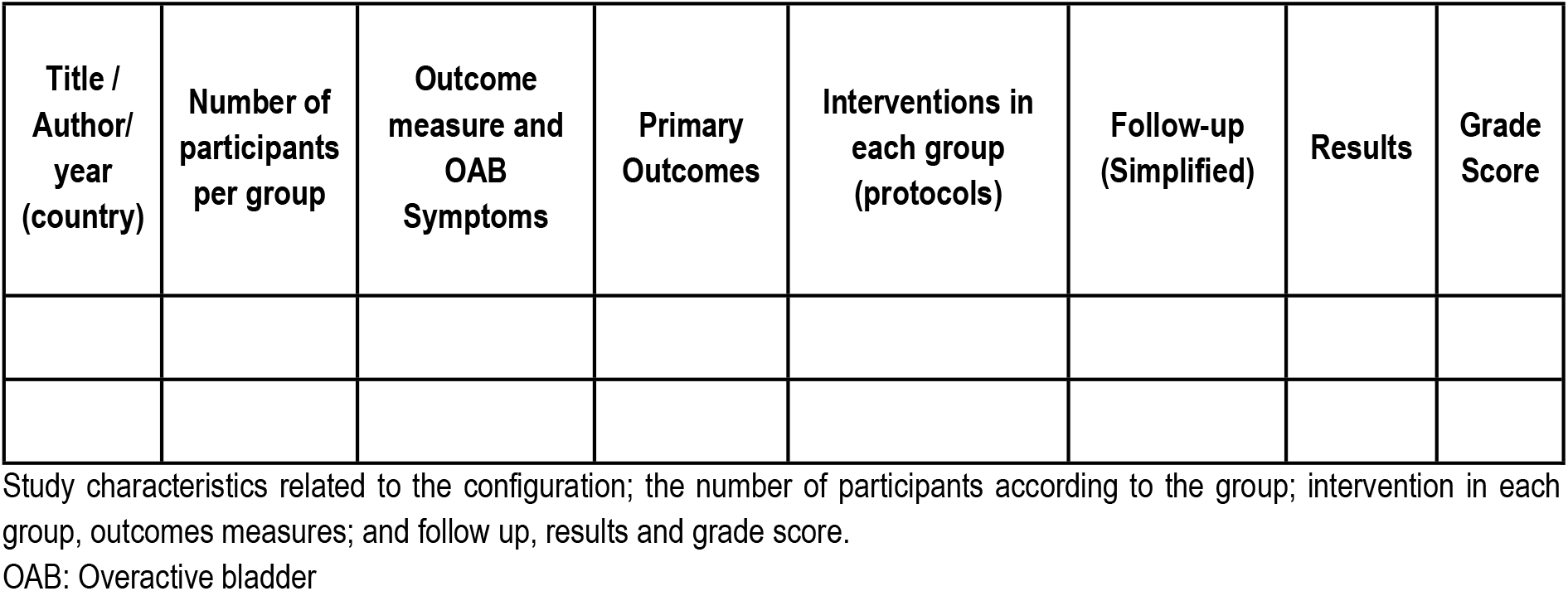
Characteristics of the Studies.

Studies that do not meet the exclusion criteria will be excluded and added in Table 2 with reason.

**Table 2.**
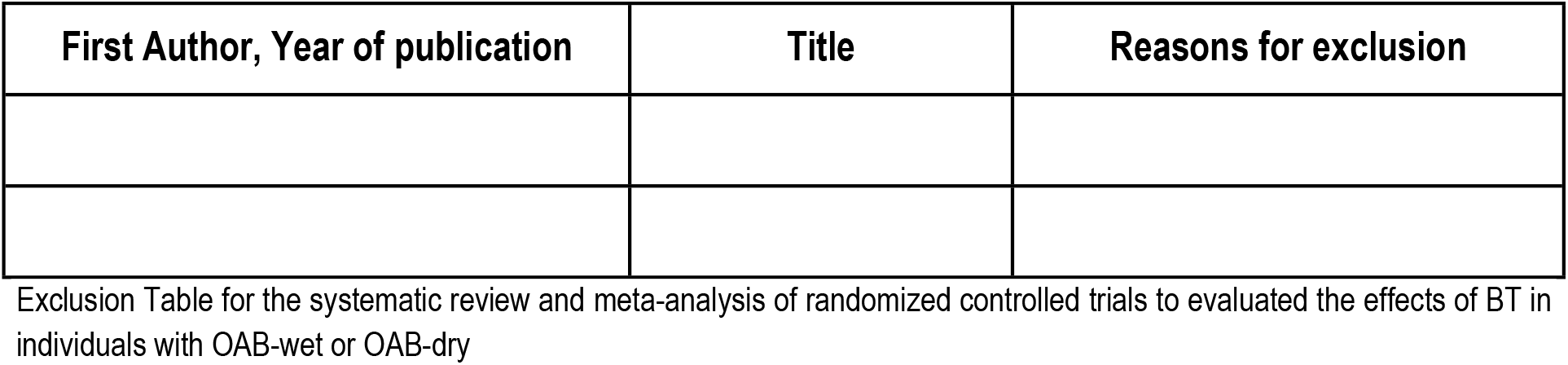
Exclusion criteria from systematic review and meta-analysis of randomized controlled trials.

## Methodological quality, risk of bias and statistical report

At this study will be measure the methodological quality the risk of bias of Cochrane Handbook of Interventions Systematic Reviews will be used, which assesses the following domains: allocation generation; concealment of allocation; blinding (of participants and researchers) and blinding of outcome assessment; the presence of incomplete data; reporting bias of information and other types of bias. The results to these domains may be “High”, “Low” or “Uncertain”. The final grade of the study will be based on the responses given to the first three domains and will be classified as having high, low or uncertain risk of bias. We will provide summaries of intervention effects for each study, calculating ratios (for dichotomous outcomes) or standardized mean differences (for continuous outcomes). Therefore, if possible, due to the range of different outcomes measured across the small number of existing trials, we will conduct a meta-analysis. However, if it is possible to have a study with the same type of intervention, a comparator, and the same outcome measure, we can reproduce a random-effects meta-analysis with standardized mean differences outcomes and risk ratios for binary outcomes. Ninety-five percent (95%) confidence intervals and two-sided p-values for each outcome. For verify the heterogeneity between studies, on effect measures will be evaluated using both the X2 test and the I^2^; statistic. We will consider a value of I^2^; greater than 50% an indicative of heterogeneity satisfactory. Will be a sensitivity analyses based on the quality of the studies. Stratified meta-analyses will be used to explore estimates of heterogeneity according to: study quality, populations studied, logistics of intervention, and content of the intervention. Review Manager software (RevMan) will be used for all analyses, including meta-analysis, if possible. [24-27].

### Quality of evidence

The Grading of Recommendations Assessment, Development and Evaluation (GRADE) will be used to verify the quality of evidence of the analyzed studies. The GRADE system assesses the limitations of the study, inconsistencies, indirect evidence, inaccuracies, and publication biases, classifying the level of evidence of the reviewed studies as high, moderate, low, or very low.

## Discussion

First-line conservative interventions for OAB-wet or OAB-dry may generate benefits for individuals with this syndrome. The BT may show and have been showing positive results due to its low complexity requiring simple technology, also because it involves behavioral changes in relation to voiding behavior and health habits, and finally due to its low cost for its effectiveness in prescription and treatment [5,16-20].

This study design was chosen because in this way, we can evaluate and update the highest level of available evidence. However, more conclusive and updated findings can be obtained that support clinical practice, in addition to promoting other studies of superior quality on this subject [23].

In this way, we can obtain solid and conclusive evidence, whether or not there is evidence to support clinical practice, in addition to promoting high-quality studies on the subject.

## Data Availability

All relevant data from this study will be made available upon study completion.

## Author Contributions

### Conceptualization

Anna Karoline Lopes Rocha, Silvia Elizate Monteiro, Maria Volpato. Dinah Verleun, Lilian Valim, Cássio Riccetto, Simone Botelho.

### Data curation

Anna Karoline Lopes Rocha, Maria Volpato, Cássio Riccetto, Simone Botelho.

### Formal analysis

Anna Karoline Lopes Rocha, Maria Volpato, Cássio Riccetto, Simone Botelho.

### Funding acquisition

Anna Karoline Lopes Rocha, Silvia Elizate Monteiro, Maria Volpato, Cássio Riccetto, Simone Botelho.

### Investigation

Anna Karoline Lopes Rocha, Dinah Verleun, Lilian Valim.

### Methodology

Anna Karoline Lopes Rocha, Silvia Elizate Monteiro, Dinah Verleun, Lilian Valim, Cássio Riccetto, Simone Botelho.

### Project administration

Cássio Riccetto, Simone Botelho.

### Resources

Anna Karoline Lopes Rocha, Dinah Verleun, Lilian Valim.

### Software

Anna Karoline Lopes Rocha, Dinah Verleun, Maria Volpato

### Supervision

Silvia Elizate Monteiro, Cássio Riccetto, Simone Botelho.

### Validation

Silvia Elizate Monteiro, Lilian Valim

### Writing – original draft

Anna Karoline Lopes Rocha, Silvia Elizate Monteiro, Maria Volpato. Dinah Verleun, Lilian Valim, Cássio Riccetto, Simone Botelho.

### Writing – review & editing

Anna Karoline Lopes Rocha, Silvia Elizate Monteiro, Maria Volpato. Dinah Verleun, Lilian Valim, Cássio Riccetto, Simone Botelho.

## Acknowledgements

We thank you the groups of the all universities - UNIFAL-MG, UNICAMP-SP and PUC-MG as well as the *Coordenação de Aperfeiçoamento de Pessoal de Nível Superior* - Brazil (CAPES), Fundação de Amparo à Pesquisa do Estado de Minas Gerais (FAPEMIG) and Research Incentive Fund, PUC MINAS, MG, Brazil.

## Supporting Information

### S1 Table

**Table 1. Characteristics of the Studies**. OAB: Overactive Bladder

### S2 Table

**Table 2. Exclusion criteria from systematic review and meta-analysis of randomized controlled trials**. Exclusion table for the systematic review and meta-analysis of randomized controlled trials to evaluated the effects of BT in individuals with OAB-wet or OAB-dry.

### S1 File

**PRISMA-P (Preferred Reporting Items for Systematic review and Meta-Analysis Protocols) 2015 checklist: Recommended items to address in a systematic review protocol\***.

### S2 File

**PROSPERO International Prospective Register of Systematic Reviews**.

## Abbreviations

ICS: International Continence Society
BT: Bladder Training
OAB: Overactive Bladder
OAB-wet: Overactive bladder with urinary incontinence
OAB-dry: Overactive bladder without urinary incontinence
UUI: Urgency Urinary Incontinence
RCTs: Randomized Clinical Trials
PFMT: Pelvic Floor Muscle Training
PFM: Pelvic Floor Muscle

